# Manifestation-Guided Adaptation of CT Foundation Model for Early Neurological Prognosis after ECPR

**DOI:** 10.64898/2026.11.15.26368165

**Authors:** Ardavan Kaviany, Pornthira Mutirangura, Samarth Goyal, Oladi Bentho, Andrea Elliott, Mahdi Bayat

## Abstract

**Purpose:** To develop a data-efficient, clinically guided transfer-learning framework for hyperacute neurological outcome prediction from head CT in patients undergoing extracorporeal cardiopulmonary resuscitation (ECPR) after cardiac arrest by leveraging free-text radiology report.

**Methods:** This retrospective study included 683 patients with available CT examination and radiology reports from December 2015 to November 2025. The primary outcome distinguished favorable neurological outcomes (Cerebral Performance Category [CPC] 1-2) from neurological death due to brain death or cerebral herniation (CPC 5). A cross-attention adaptor jointly learned to predict neurological outcome and five report-derived manifestations of brain injury from a frozen pretrained head CT encoder. Outcome prediction used CT images alone at inference. The manifestation-guided adaptor, trained with and without additional patients who contributed only manifestation labels, was compared with outcome-only adaptor and two pooled-feature baselines. Performance was summarized as mean ± SD across five held-out folds.

**Results:** Of 683 patients, 197 were eligible for outcome evaluation (63 with favorable outcomes and 134 with neurological death), and 486 formed the manifestation-only cohort. The area under the receiver operating characteristic curve (AUROC) was 0.918 ± 0.044 for the outcome-only adaptor. The manifestation-guided adaptor achieved an AUROC of 0.927 ± 0.042. With additional manifestation-only training data, the adaptor achieved the mean AUROC (0.929 ± 0.037), specificity (93.7% ± 6.5%), with a sensitivity of 79.9% ± 9.2%.

**Conclusion:** Manifestation-guided adaptation outperforms outcome-only adaptation for neurological outcome prediction from hyperacute head CT after ECPR. Report-derived manifestation labels enabled training with additional examinations without eligible outcome labels while preserving CT-only prediction.

## Introduction

Hypoxic-ischemic brain injury (HIBI) is among the most consequential complications of cardiac arrest and a major determinant of neurological outcome after resuscitation (1). Because injury evolves over hours and days, its eventual severity is difficult to assess during the earliest phase of care. Guidelines recommend multimodal neuroprognostication, with impressions delayed until potential confounders, such as residual sedation, temperature-related effects, and organ dysfunction, have been minimized (1). Early neurological risk stratification can prioritize lifesaving care to patients who warrant closer monitoring and further diagnostic evaluation. The Extracorporeal Life Support Organization recommends CT as soon as practical after extracorporeal cardiopulmonary resuscitation (ECPR) to identify catastrophic brain injury and other complications (2).

Noncontrast head CT provides an accessible means of evaluating HIBI. Cerebral edema, loss of gray-white matter differentiation, and sulcal effacement have been associated with neurological outcomes after cardiac arrest, including in patients undergoing ECPR (3,4). Early CT findings can also influence treatment decisions. Beekman et al. reported that abnormal findings on head CT obtained within six hours after cardiac arrest prompted changes in care, most commonly treatment limitation or withdrawal of life-sustaining treatment (5). However, early CT features may be subtle and incompletely developed, and the sensitivity of quantitative gray-white matter assessment varies with imaging timing (6). Previous studies have investigated radiomics, transfer learning, and multimodal models combining CT with clinical data for early outcome prediction (7–9). These approaches support the potential of CT-based prediction, although limited outcome-labeled data remain a challenge for model development. Although markedly reduced gray-white matter ratio can be highly specific for poor neurological outcomes, conventional CT markers have shown limited sensitivity and lower discriminative performance within the first six hours after cardiac arrest compared with later imaging (6,10,11).

Self-supervised pretraining can support medical image classification with fewer labeled examples (12). Radiology reports provide a complementary source of supervision by documenting clinically meaningful imaging findings. Report-derived multitask supervision has improved imaging representations for downstream mortality prediction, supporting its potential use beyond abnormality detection (13). Such supervision could also permit training with examinations lacking outcome labels suitable for the primary prediction task. However, whether report-derived manifestations improve adaptation of pretrained head CT representations for hyperacute outcome prediction after ECPR remains unclear. We hypothesized that manifestation-guided adaptation would improve prediction performance when outcome-labeled training data are limited. Therefore, this study aimed to develop a data-efficient, clinically guided transfer-learning framework for hyperacute neurological outcome prediction from head CT obtained within six hours after cannulation in patients undergoing ECPR after cardiac arrest using free-text radiology reports.

## Materials and Methods

### Study Design and Data

This single-center retrospective study included consecutive patients who underwent ECPR after cardiac arrest at the [University of Minnesota] between December 2015 to November 2025 (n = 1321). The Study was approved by the institutional review board, and the requirement for informed consent was waived. Patients who had declined research authorization were excluded. Eligible examinations were non-contrast head CT scans acquired within 6 hours after ECPR cannulation with corresponding free-text radiology reports. The final study cohort included 683 patients, with one eligible examination per patient (Fig 1). Clinicians assigned discharge Cerebral Performance Category (CPC) scores and determined cause of death based on chart-reviewed clinical information. The primary prediction task distinguished favorable neurological outcome (CPC 1–2) from catastrophic neurological outcome, defined as CPC 5 attributable to brain death or cerebral herniation. These endpoints were selected to focus the analysis on favorable recovery versus fatal neurological injury, consistent with the intended application of supporting early neurological risk assessment. Patients with CPC 3–4, unknown discharge outcomes, or deaths associated with withdrawal of life-sustaining treatment or non-neurological causes were excluded from the primary outcome analysis. The primary outcome cohort comprised 197 patients, including 63 with favorable outcomes and 134 with catastrophic neurological outcomes. The remaining 486 patients contributed report-derived manifestation supervision during model development but were not included in the primary outcome evaluation.

**Figure 1.**
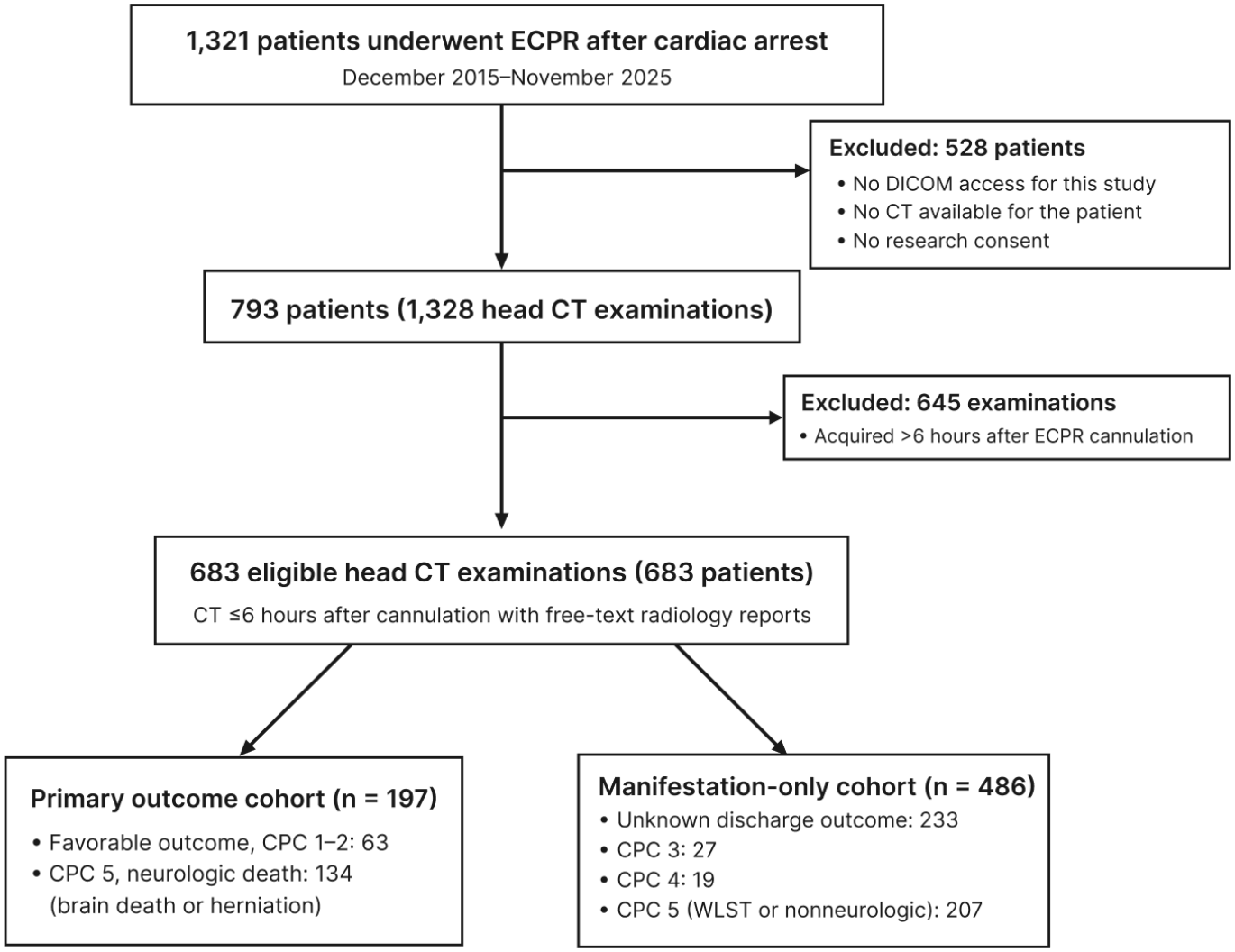
Flowchart shows the patient selection for the final study dataset: 197 patients in the primary outcome cohort and 486 patients in the manifestation-only cohort

## Model

We developed a manifestation-guided transfer-learning framework to predict neurological outcome from hyperacute head CT (Fig 2). The framework adapted representations from a pretrained head CT encoder using neurological outcome labels, with report-derived manifestation labels providing auxiliary supervision during training. At inference, outcome prediction required only CT images.

**Figure 2.**
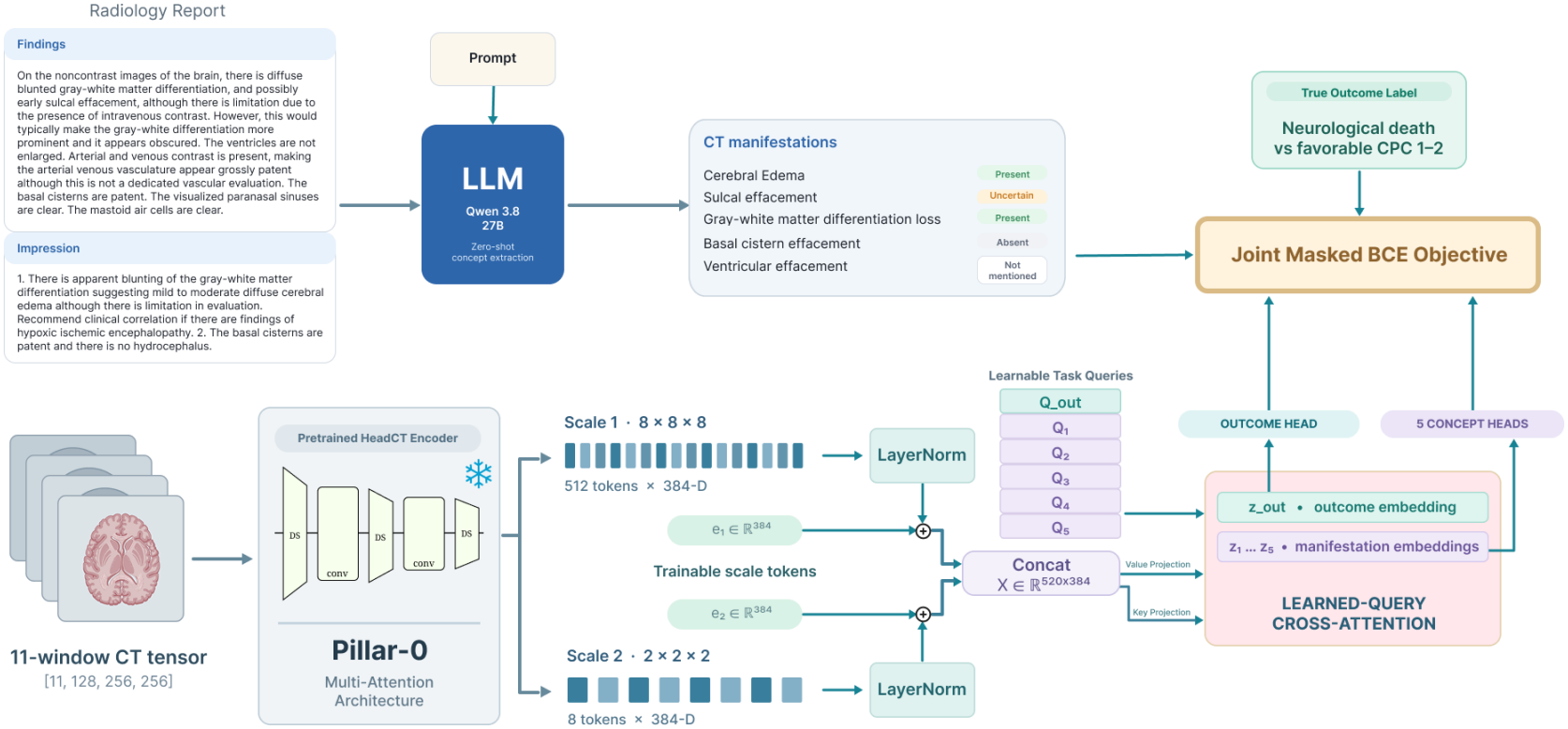
Manifestation-guided framework for neurological outcome prediction from hyperacute head CT. In the upper pathway, a language model uses a structured extraction prompt to assign report-derived labels to five CT manifestations. In the lower pathway, the frozen Pillar-0 encoder produces 512 regional and eight coarse spatial tokens. After layer normalization and addition of learned scale embeddings, the tokens are concatenated and projected into keys and values shared across tasks. Within the cross-attention adaptor, six learned queries attend to the keys and aggregate the values into one outcome representation and five manifestation representations, each connected to a separate prediction head. The adaptor and prediction heads are trained jointly using weighted, masked losses based on available outcome and manifestation labels. Uncertain and unmentioned manifestations and unavailable outcome labels do not contribute to their respective losses. Radiology reports provide training supervision; outcome prediction at inference requires only CT images.

Five CT manifestations of hypoxic-ischemic brain injury were selected as auxiliary targets with clinician approval: loss of gray-white differentiation, cerebral edema or swelling, sulcal effacement, basal cistern effacement, and ventricular effacement. These findings reflect changes in brain parenchymal attenuation and the anatomical effects of cerebral swelling and have been described in previous studies of post–cardiac arrest CT, including analyses of radiology reports (14,15). Qwen3.8-27B (16) was used for zero-shot structured extraction with greedy decoding. The prompt specified manifestation definitions, labeling rules, and an output schema to classify each manifestation as present, absent, uncertain, or not mentioned.

Head CT examinations were preprocessed using the RAVE preprocessing engine (17) and passed through the pretrained Pillar-0 encoder (17), whose parameters remained frozen during training. After layer normalization, learned scale embeddings were added to the regional and coarse spatial tokens to distinguish the two feature scales. The tokens were then concatenated and supplied to a cross-attention adaptor with six learned task-specific queries: one for neurological outcome and five for the manifestations. Each query attended to the shared image features and aggregated them into a separate representation, allowing spatial information to be weighted differently for each task. Each representation passed to an independent prediction head to estimate the probability of neurological outcome or the corresponding manifestation.

The cross-attention adaptor and prediction heads were trained jointly using a weighted combination of outcome and manifestation binary cross-entropy losses. Manifestation supervision updated shared adaptor parameters used to construct the outcome representation, while the outcome head received only its dedicated representation. Explicitly present and absent manifestations were encoded as positive and negative targets, respectively; uncertain and unmentioned findings were masked from the corresponding losses. Outcome loss was calculated only for patients with eligible outcome labels, allowing the remaining patients to contribute manifestation supervision alone.

### Model Evaluation and Outcome Prediction

The primary analysis evaluated the ability of the models to predict neurological outcome using the area under the receiver operating characteristic curve (AUROC). Neurological death was designated as the positive class. The area under the precision-recall curve (AUPRC) and threshold-dependent classification metrics were reported as secondary performance measures.

To assess the contribution of spatial adaptation, an outcome-only cross-attention adaptor was compared with logistic regression and a MLP using pooled representations from the same frozen encoder. The contribution of manifestation supervision was assessed by comparing outcome-only and manifestation-guided adaptation within the same outcome-eligible cohort. The contribution of additional report-derived supervision was assessed by comparing manifestation-guided adaptation with and without the cohort of 486 manifestation-only patients.

Fivefold patient-level cross-validation was performed. Outcome-eligible patients were stratified by neurological outcome, and manifestation-only patients were stratified separately by manifestation burden and assigned to the same five outer folds, so each held-out fold contained patients from both cohorts.

Within each outer training set, patients were divided into inner training and validation partitions comprising approximately 80% and 20%, respectively, with manifestation-only patients retained as a separate stratum. Among manifestation-only patients, only those assigned to the inner training partition contributed supervision. Model checkpoints were selected by minimizing outcome binary cross-entropy calculated exclusively on outcome-eligible patients in the inner validation partition. The manifestation-supervision weight was selected independently within each outer fold from 0.1, 0.25, 0.5, 1.0, and 2.0 by maximizing outcome AUROC on these inner-validation patients. Neurological outcome prediction performance was evaluated separately within each held-out outer fold using patients with eligible outcome labels.

Sensitivity, specificity, positive and negative predictive values, accuracy, balanced accuracy, and F1 score were calculated using thresholds selected by maximizing Youden’s J statistic on the inner validation partition. For each model and outer fold, the selected threshold was applied to the corresponding held-out predictions without further adjustment.

Manifestation prediction was evaluated using held-out examinations from both cohorts, restricted for each manifestation to scans with explicitly present or absent report-derived labels. AUROC, AUPRC, and threshold-dependent classification metrics were calculated separately for each manifestation within each outer test fold. Manifestation-specific thresholds were selected by maximizing Youden’s J statistic on explicitly labeled examinations in the corresponding inner validation partition and applied to that fold’s held-out predictions without adjustment. For each metric, macro-averaged performance was calculated within each outer test fold as the unweighted mean across the five manifestations. These analyses assessed agreement between model predictions and findings documented in radiology reports.

To assess data efficiency, the MLP baseline and cross-attention adaptors were retrained using 25%, 50%, and 75% of the inner-training patients, while validation and test partitions remained fixed. At each fraction, five independent stratified subsamples were generated and shared across models. For models incorporating manifestation-only patients, this additional training cohort was reduced proportionally.

Manifestation-loss weights were retained from the full-data analysis. For each reduced training fraction, pooled outer-test AUROC was calculated separately for each of the five repeated subsamples and summarized as the mean and standard deviation across repetitions. The result at 100% was obtained from the primary cross-validation analysis without retraining.

For the primary neurological outcome and manifestation prediction analyses, performance metrics were summarized as the arithmetic mean ± standard deviation across the five held-out outer folds.

Macro-averaged manifestation performance was summarized using the five fold-specific macro-averages. Model comparisons were descriptive, based on mean performance differences across the same outer folds. Out-of-fold image representations from the frozen encoder and the adapted models were visualized using Uniform Manifold Approximation and Projection (UMAP). Local outcome-label agreement was assessed using 10-nearest-neighbor (10-NN) purity, defined as the proportion of neighboring outcome-eligible patients sharing the same neurological outcome. Patients in the manifestation-only cohort were included in the visualization but excluded from the neighborhood-purity calculation.

Outcome-query attention maps were generated using the manifestation-guided adaptor trained with all patients. Each patient was visualized using their held-out outer-fold model. Softmax cross-attention weights for the 512 regional tokens were mapped to an 8×8×8 grid.

## Data and Code Availability

The imaging, radiology report, and clinical data used in this study are not publicly accessible to ensure patient privacy protection. The code for data preprocessing, report-label extraction, model training, and evaluation are available at https://github.com/AKV002/GuidedAdaptation_EarlyCT

## Results

### Dataset Characteristics

A total of 683 patients with 683 eligible head CT examinations were included in the study cohort (mean age ± SD, 57.3 ± 13.1 years; 178 female patients [26.1%]). The median interval from ECPR cannulation to CT was 2.08 hours (IQR, 1.57–2.78 hours). The primary outcome cohort comprised 197 patients, including 63 (32.0%) with favorable neurological outcomes and 134 (68.0%) with neurological death. The remaining 486 patients constituted the manifestation-only cohort. Patient characteristics are summarized in Table 1.

**Table 1.**
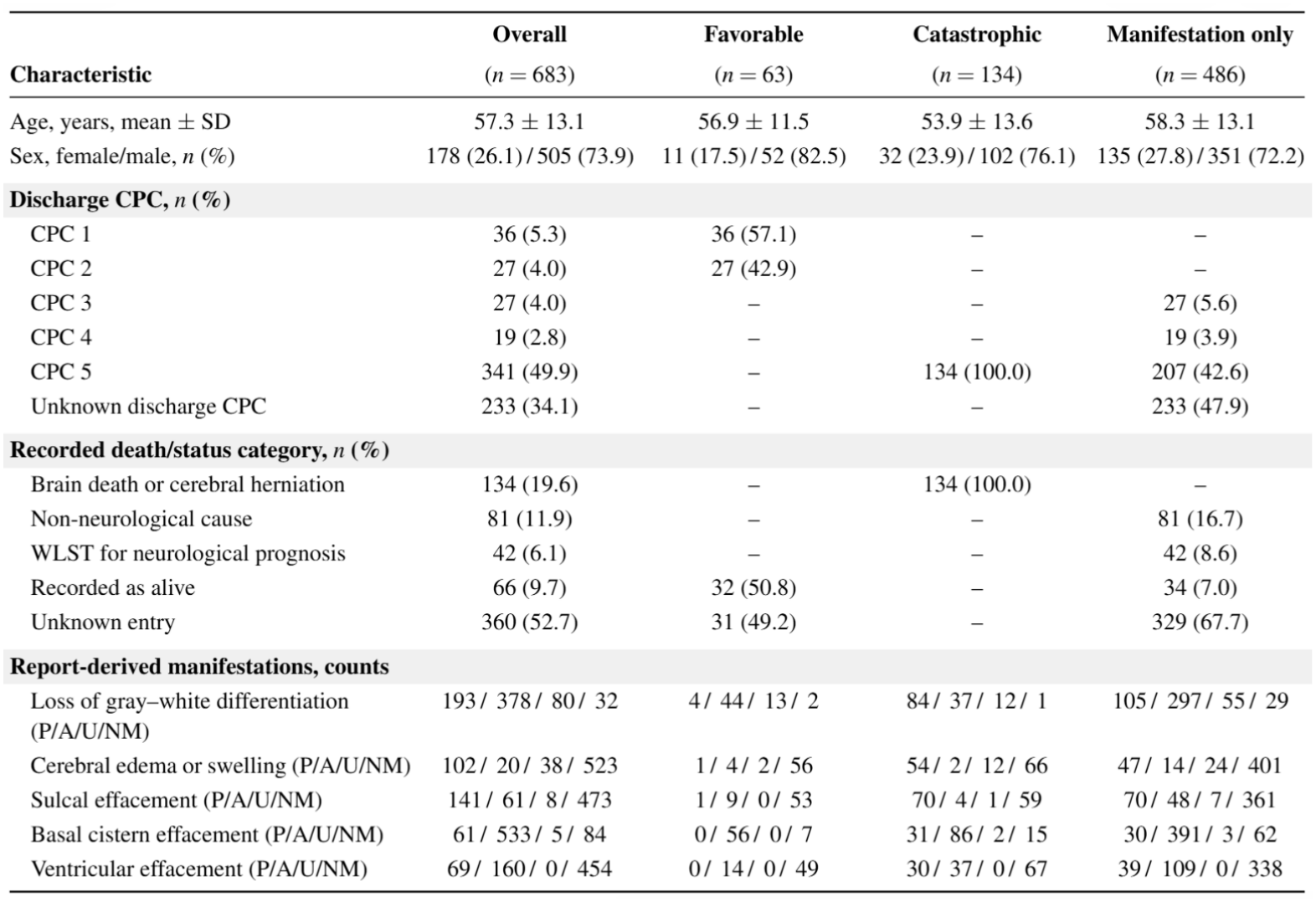
Patient characteristics, neurological outcomes, and report-derived CT manifestations. One CT examination was included per patient. Manifestation counts are reported as present/absent/uncertain/not mentioned (P/A/U/NM). WLST = withdrawal of life-sustaining treatment.

|  | **Overall** | **Favorable** | **Catastrophic** | **Manifestation only** |
|---|---|---|---|---|
| **Characteristic** | (*n* = 683) | (*n* = 63) | (*n* = 134) | (*n* = 486) |
| Age, years, mean ± SD | 57.3 ± 13.1 | 56.9 ± 11.5 | 53.9 ± 13.6 | 58.3 ± 13.1 |
| Sex, female/male, *n* (%) | 178 (26.1) / 505 (73.9) | 11 (17.5) / 52 (82.5) | 32 (23.9) / 102 (76.1) | 135 (27.8) / 351 (72.2) |
| **Discharge CPC, *n* (%)** |  |  |  |  |
| CPC 1 | 36 (5.3) | 36 (57.1) | – | – |
| CPC 2 | 27 (4.0) | 27 (42.9) | – | – |
| CPC 3 | 27 (4.0) | – | – | 27 (5.6) |
| CPC 4 | 19 (2.8) | – | – | 19 (3.9) |
| CPC 5 | 341 (49.9) | – | 134 (100.0) | 207 (42.6) |
| Unknown discharge CPC | 233 (34.1) | – | – | 233 (47.9) |
| **Recorded death/status category, *n* (%)** |  |  |  |  |
| Brain death or cerebral herniation | 134 (19.6) | – | 134 (100.0) | – |
| Non-neurological cause | 81 (11.9) | – | – | 81 (16.7) |
| WLST for neurological prognosis | 42 (6.1) | – | – | 42 (8.6) |
| Recorded as alive | 66 (9.7) | 32 (50.8) | – | 34 (7.0) |
| Unknown entry | 360 (52.7) | 31 (49.2) | – | 329 (67.7) |
| **Report-derived manifestations, counts** |  |  |  |  |
| Loss of gray–white differentiation (P/A/U/NM) | 193 / 378 / 80 / 32 | 4 / 44 / 13 / 2 | 84 / 37 / 12 / 1 | 105 / 297 / 55 / 29 |
| Cerebral edema or swelling (P/A/U/NM) | 102 / 20 / 38 / 523 | 1 / 4 / 2 / 56 | 54 / 2 / 12 / 66 | 47 / 14 / 24 / 401 |
| Sulcal effacement (P/A/U/NM) | 141 / 61 / 8 / 473 | 1 / 9 / 0 / 53 | 70 / 4 / 1 / 59 | 70 / 48 / 7 / 361 |
| Basal cistern effacement (P/A/U/NM) | 61 / 533 / 5 / 84 | 0 / 56 / 0 / 7 | 31 / 86 / 2 / 15 | 30 / 391 / 3 / 62 |
| Ventricular effacement (P/A/U/NM) | 69 / 160 / 0 / 454 | 0 / 14 / 0 / 49 | 30 / 37 / 0 / 67 | 39 / 109 / 0 / 338 |

Across the five manifestations, radiology reports provided 1718 explicitly present or absent labels, including 568 from the primary outcome cohort and 1150 from the manifestation-only cohort. Label availability varied by manifestation, ranging from 122 of 683 patients (17.9%) for cerebral edema or swelling to 594 of 683 patients (87.0%) for basal cistern effacement. Within the primary outcome cohort, the number of patients with evaluable report-derived labels ranged from 61 to 173 per manifestation.

## Model Prediction Performance

Across five fold patient-level cross-validation in 197 outcome-eligible patients, mean AUROCs ranged from 0.891 to 0.929 (Table 2). The pooled-feature logistic regression and multilayer perceptron achieved AUROCs of 0.894 ± 0.045 and 0.891 ± 0.043, respectively (mean ± SD). The outcome-only adaptor achieved a higher AUROC of 0.918 ± 0.044, with an AUPRC of 0.964 ± 0.019 compared with 0.954 ± 0.022 and 0.951 ± 0.026 for the respective pooled-feature baselines. ROC curves for the multilayer perceptron, outcome-only adaptor, and manifestation-guided adaptor are shown in Figure 3A.

**Table 2.**
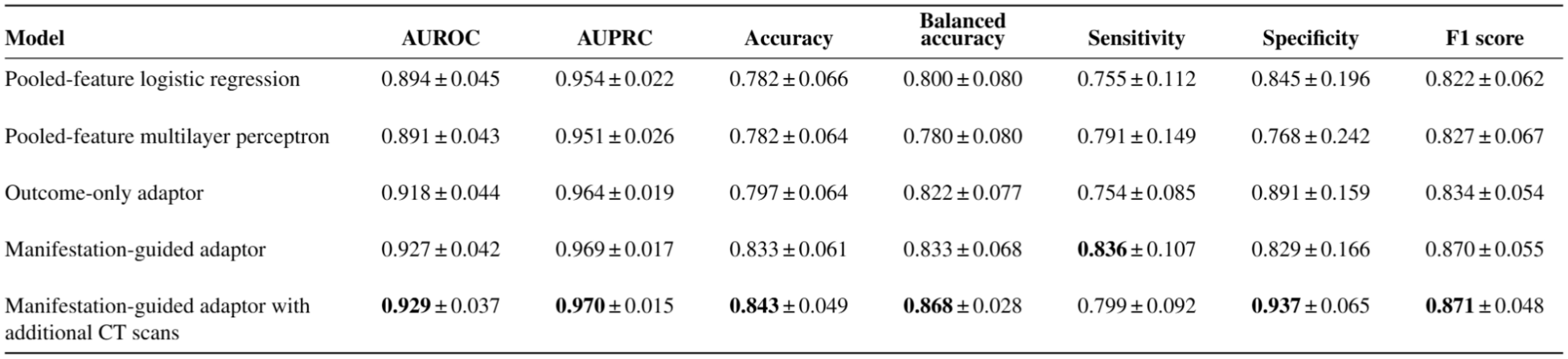
Neurological Outcome Prediction result.

**Figure 3.**
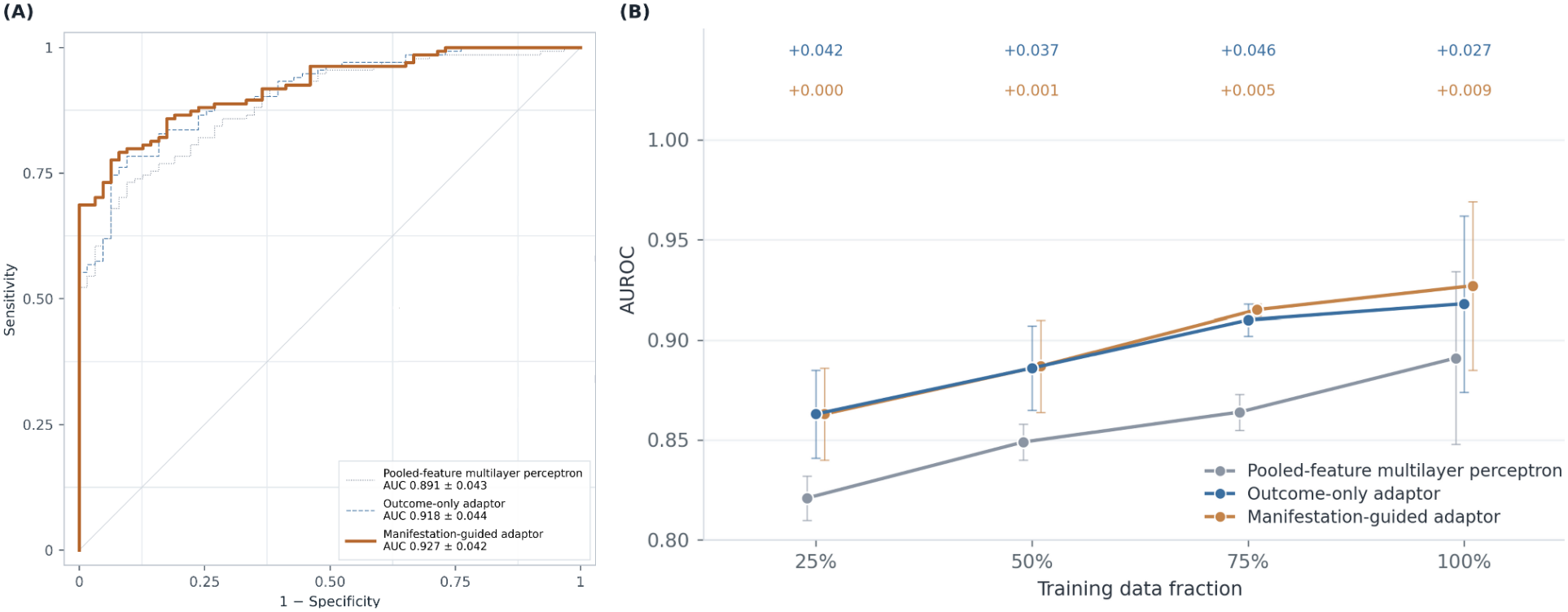
Discrimination and data efficiency of head CT–based neurological outcome prediction models. (A) Receiver operating characteristic curves comparing the pooled-feature MLP, outcome-only adaptor, and manifestation-guided adaptor. (B) AUROC using 25%, 50%, 75%, and 100% of the available training data. Points represent mean performance, and error bars indicate SD. Blue annotations show the absolute AUROC gain of the outcome-only adaptor over the pooled-feature MLP. Orange annotations show the additional gain from manifestation guidance.

Within the same outcome-eligible cohort, manifestation-guided adaptation achieved an AUROC of 0.927 ± 0.042 and an AUPRC of 0.969 ± 0.017, compared with 0.918 ± 0.044 and 0.964 ± 0.019 for outcome-only adaptation. At the fold-specific thresholds selected in the inner validation partitions, manifestation-guided adaptation had higher mean sensitivity (83.6% vs 75.4%) but lower mean specificity (82.9% vs 89.1%). Mean accuracy was 83.3% versus 79.7%, and mean balanced accuracy was 83.3% versus 82.2%.

Incorporating report-derived supervision from the additional manifestation-only cohort yielded the highest observed mean AUROC (0.929 ± 0.037) and AUPRC (0.970 ± 0.015), representing increments of 0.002 and 0.001 over manifestation-guided adaptation without additional scans. Mean specificity increased from 82.9% to 93.7%, while mean sensitivity decreased from 83.6% to 79.9%. Mean balanced accuracy increased from 83.3% to 86.8%, and mean accuracy increased from 83.3% to 84.3%. Compared with the outcome-only adaptor, the model incorporating additional scans had both higher mean sensitivity (79.9% vs 75.4%) and higher mean specificity (93.7% vs 89.1%).

In the data-efficient training analyses, the outcome-only adaptor achieved higher mean AUROC than the multilayer perceptron at the 25%, 50%, and 75% training fractions, with absolute differences of 0.042, 0.037, and 0.046, respectively (Fig 3B). The corresponding AUROC differences between manifestation-guided and outcome-only adaptation were 0.000, 0.001, and 0.005. At the full training fraction, the AUROC difference between outcome-only adaptation and the multilayer perceptron was 0.027, while manifestation guidance provided an additional difference of 0.009.

UMAP visualization of outcome representations showed increased local outcome-label agreement following manifestation-guided adaptation, as reflected by higher 10-NN purity, with further improvement when additional manifestation-only scans were incorporated (Fig 4). These findings complement the improvements in neurological outcome discrimination observed in the primary performance analysis.

**Figure 4.**
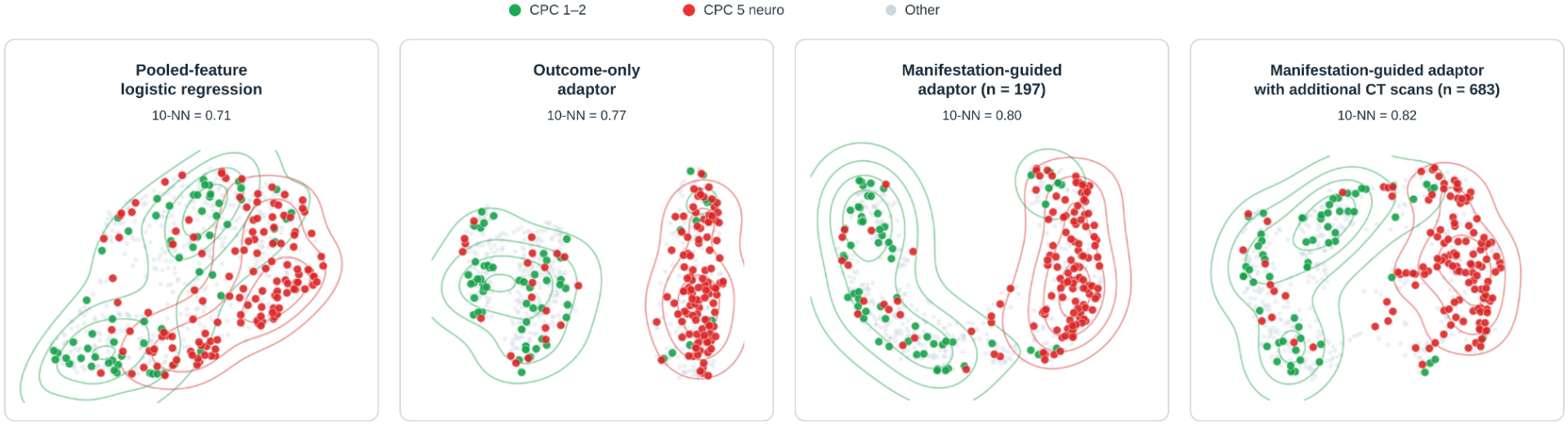
UMAP visualizations compare outcome-only and manifestation-guided adaptation with and without additional CT scans, with points colored by neurological outcome and 2D 10-NN purity reported for each representation.

The manifestation-guided adaptor incorporating additional CT scans achieved a macro-averaged AUROC of 0.921 ± 0.012 for manifestation classification (Table 3). AUROCs ranged from 0.881 ± 0.008 for gray-white differentiation loss to 0.958 ± 0.022 for basal cistern effacement, with corresponding values of 0.951 ± 0.062 for cerebral edema and 0.930 ± 0.045 for sulcal effacement. Macro-averaged balanced accuracy and F1 score were 0.807 ± 0.013 and 0.739 ± 0.054, respectively. Incorporating additional report-derived supervision during training yielded higher mean AUROCs for all five manifestations, with an absolute increase of 0.010 in macro-averaged AUROC. Outcome-query attention maps for four representative patients are shown in Figure 5.

**Table 3.**
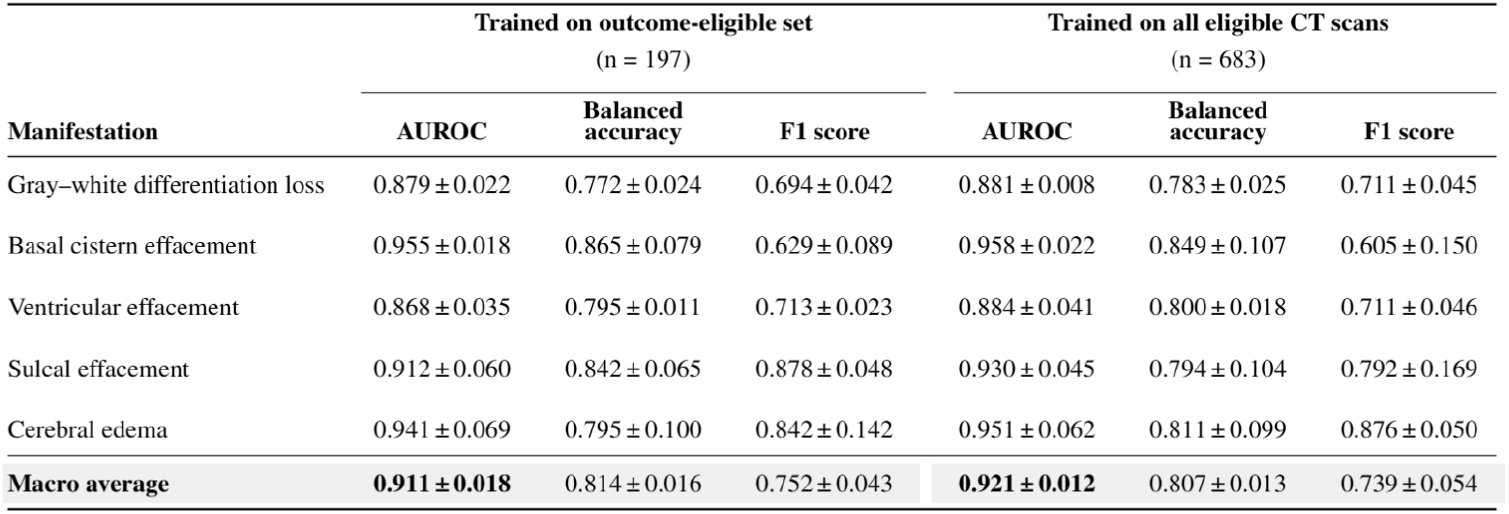
CT Manifestation Classification result.

| Manifestation | Trained on outcome-eligible set<br>(n = 197) |  |  | Trained on all eligible CT scans<br>(n = 683) |  |  |
|---|---|---|---|---|---|---|
|  | **AUROC** | **Balanced accuracy** | **F1 score** | **AUROC** | **Balanced accuracy** | **F1 score** |
| Gray–white differentiation loss | 0.879 ± 0.022 | 0.772 ± 0.024 | 0.694 ± 0.042 | 0.881 ± 0.008 | 0.783 ± 0.025 | 0.711 ± 0.045 |
| Basal cistern effacement | 0.955 ± 0.018 | 0.865 ± 0.079 | 0.629 ± 0.089 | 0.958 ± 0.022 | 0.849 ± 0.107 | 0.605 ± 0.150 |
| Ventricular effacement | 0.868 ± 0.035 | 0.795 ± 0.011 | 0.713 ± 0.023 | 0.884 ± 0.041 | 0.800 ± 0.018 | 0.711 ± 0.046 |
| Sulcal effacement | 0.912 ± 0.060 | 0.842 ± 0.065 | 0.878 ± 0.048 | 0.930 ± 0.045 | 0.794 ± 0.104 | 0.792 ± 0.169 |
| Cerebral edema | 0.941 ± 0.069 | 0.795 ± 0.100 | 0.842 ± 0.142 | 0.951 ± 0.062 | 0.811 ± 0.099 | 0.876 ± 0.050 |
| **Macro average** | **0.911 ± 0.018** | 0.814 ± 0.016 | 0.752 ± 0.043 | **0.921 ± 0.012** | 0.807 ± 0.013 | 0.739 ± 0.054 |

**Figure 5.**
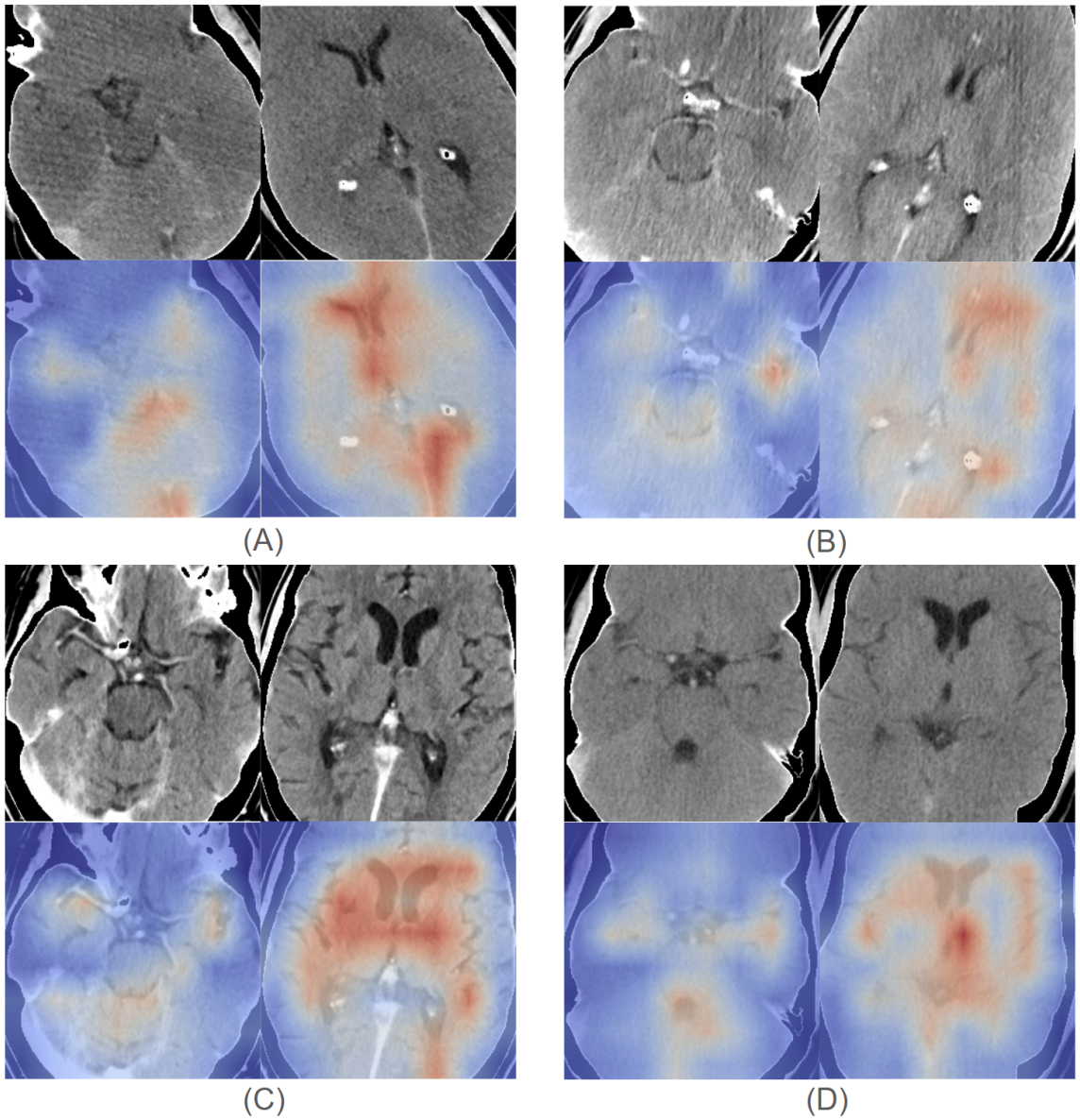
Regional outcome-query attention across differing CT manifestations and neurological outcomes. Axial CT images (upper rows) and corresponding attention overlays (lower rows) are shown for four correctly classified patients. Warmer colors indicate higher relative attention weights. **(A)** Neurological death with all five manifestations reported and predicted, with attention involving anterior and posterior periventricular regions. **(B)** Neurological death with no manifestations reported on the initial CT, although the model predicted cerebral edema, basal cistern effacement, and ventricular effacement. Attention involves periventricular and adjacent deep cerebral regions. All five manifestations were reported on subsequent examinations. (C) Favorable outcome with reported mild loss of gray-white differentiation and a corresponding positive manifestation prediction, accompanied by broad bilateral periventricular and deep gray attention. **(D)** Favorable outcome without reported or predicted manifestations, with attention involving central deep gray regions. Higher attention does not directly indicate greater injury severity or establish whether regional features increased or decreased the predicted probability of neurological death.

## Discussion

Manifestation-guided adaptation offers a practical approach to neurological outcome prediction from early head CT when outcome-labeled data are limited. The adaptor trained with additional CT scans achieved a mean AUROC of 0.929 ± 0.037, compared with 0.918 ± 0.044 for the outcome-only adaptor. It also classified five manifestations of brain injury with a macro-average AUROC of 0.921 ± 0.012. These findings suggest that routinely available radiology reports can provide useful auxiliary supervision, enabling patients outside the outcome-eligible cohort to contribute to model training while preserving CT-only inference.

Previous work has shown that large-scale pretraining can improve medical image classification when labeled data are limited (12). Eyuboglu et al. further demonstrated that representations learned through report-derived abnormality supervision improved subsequent mortality prediction from FDG-PET/CT (13). Our comparisons suggest that task-specific adaptation and manifestation supervision provide complementary benefits. The outcome-only adaptor achieved higher mean AUROC than both pooled-feature classifiers using the same frozen encoder, supporting the value of learning how to aggregate spatial CT features for the prediction task. Adding manifestation supervision produced a further numerical improvement within the same outcome-eligible cohort, suggesting that these labels provide useful information beyond outcome supervision alone. The data-efficiency analysis also showed an advantage for the adaptor over the pooled-feature baseline at reduced training sizes, whereas the incremental benefit of manifestation guidance became more apparent as training data increased.

The manifestation classification results indicate that the adapted CT representations contain information relevant to several features of brain injury. Discrimination was highest for basal cistern effacement and cerebral edema, with mean AUROCs of 0.958 ± 0.022 and 0.951 ± 0.062, respectively. Predicting these manifestations provides specific imaging findings that can accompany the neurological outcome estimate. Incorporating additional scans increased AUROC across all five manifestations, supporting the value of expanding report-derived supervision. Changes in balanced accuracy and F1 score were less consistent, indicating that improvements in discrimination did not uniformly translate into better classification at the selected thresholds. Outcome-query attention emphasized periventricular and deep gray regions across the representative cases (Fig 5). Overlapping anatomical territories received attention in patients with both neurological death and favorable outcomes, including a patient without reported or predicted manifestations. The favorable-outcome case with reported and predicted gray-white differentiation loss illustrates the distinction between manifestation detection and outcome prediction. For the patient shown in figure 5B, manifestations reported on subsequent CT provide context for the early model predictions but do not establish earlier detection of injury. These maps offer a qualitative view of regional feature weighting localized on anatomically meaningful parts of the brain soft tissue; future studies with extended cohorts will allow better clinical association and explainability.

The use of routine reports involved a trade-off between annotation burden and label completeness. Although this approach avoided additional image annotation, it relied on findings explicitly documented by radiologists. Masking uncertain and unmentioned findings prevented unreported manifestations from being treated as absent, but reduced usable supervision and could not resolve inaccuracies in the reports or errors during label extraction. Manifestation performance was therefore estimated in the subset with explicit labels, which may differ from examinations with uncertain or unreported findings. Independent image adjudication is needed to establish performance across the full range of manifestation presentations.

The primary outcome analysis included 197 patients from a single center because it required both head CT within six hours of ECPR cannulation and a discharge outcome that met the prespecified criteria. Internal cross-validation estimated performance within this cohort, leaving generalizability to other centers uncertain. We are unaware of a publicly available dataset with comparable CT timing and neurological outcome definitions. As more patients become available, we plan to expand the development cohort and pursue external validation with data from other ECPR centers. A second limitation was the use of CT as the sole input at inference. This design allowed us to assess whether report-derived manifestations could guide the adaptation of pretrained CT representations for neurological outcome prediction. Guidelines recommend multimodal neuroprognostication (1,18,19). Future work will integrate the adapted CT representation with electronic health record (EHR) data and findings from neurological examinations and other tests to determine what early CT adds to a multimodal assessment.

We developed a manifestation-guided framework that uses radiology reports to adapt pretrained head CT representations for early neurological outcome prediction after ECPR. The framework achieved higher mean AUROC than outcome-only adaptation and pooled-feature baselines while also classifying five manifestations of brain injury. This approach has the potential to support early neurological risk assessment and provide an imaging component for future multimodal models incorporating EHR and relevant clinical examinations.

## Data Availability

The patient-level head CT images, radiology reports, and clinical data used in this study are not publicly available due to patient privacy restrictions. Aggregate results are reported in the manuscript.

https://github.com/AKV002/GuidedAdaptation_EarlyCT

